# Towards environmental detection of Chagas disease vectors and pathogen

**DOI:** 10.1101/2021.12.24.21268369

**Authors:** Grace Gysin, Plutarco Urbano, Luke Brandner-Garrod, Shahida Begum, Mojca Kristan, Thomas Walker, Carolina Hernández, Juan David Ramírez, Louisa A. Messenger

## Abstract

**Background:** Accurate surveillance of triatomine household infestation is crucial for Chagas disease vector control. However, no gold standard detection method with high levels of sensitivity or specificity is currently available. Several intrinsic features of triatomine bug behaviour and the lifecycle of *Trypanosoma (T*.*) cruzi* lead to deposition of environmental DNA (eDNA) in infested houses. This study evaluated the use of FTA cards and cotton-tipped swabs as low-technology, cost-effective tools for simultaneous detection of *T. cruzi* and vector eDNA in the laboratory and field.

**Methods/Principal Findings:** This study had two components: (1) laboratory evaluation and optimisation of QIAcard® FTA® classic cards to detect *Rhodnius (R*.*) prolixus* eDNA by altering five different environmental variables (darkness, triatomine number, temperature, feeding status and degradation at ambient temperature); (2) detection of *R. prolixus* and *T. cruzi* eDNA from cotton-tipped house wall swabs from an endemic region in Casanare Department, Colombia. eDNA was extracted from all specimens and amplified using a multiplex TaqMan qPCR assay targeting the *R. prolixus 12S rRNA* gene and *T. cruzi* satellite DNA region. *R. prolixus* eDNA from five 3^rd^/4^th^ instar nymphs was successfully amplified from FTA cards after as little as 15 minutes of contact time under standard insectary conditions. Factors significantly increasing eDNA detection from FTA cards were increasing temperature from 21°C to 27-32°C, triatomine bug density from 1-25 bugs and recent blood-feeding. eDNA was detectable from FTA cards stored at room temperature for at least two weeks. In cotton-tipped swabs from the field, the sensitivity and specificity of *R. prolixus* eDNA detection was 60.6% (n=20/33) and 100% (n=33/33), respectively. *T. cruzi* eDNA was amplified from 93.9% (n=31/33) of infested houses.

**Conclusions/Significance:** FTA cards are a highly sensitive tool for entomological surveillance of *R. prolixus* and exhibit little variability under different environmental conditions. Additionally, cotton-tipped swabs are a relatively sensitive tool for entomological and parasitological surveillance of *R. prolixus* and *T. cruzi in situ*, but more feasible due to low cost. Both methods could be utilised by citizen science initiatives to contribute to the control of Chagas disease in endemic communities.

**Author Summary:** Chagas disease vector control relies on prompt, accurate identification of houses infested with triatomine bugs for targeted insecticide spraying. However, most current detection methods are laborious, lack standardization, have substantial operational costs and limited sensitivity, especially when triatomine bug densities are low or highly focal. We evaluated the use of FTA cards or cotton-tipped swabs to develop a low-technology, non-invasive method of detecting environmental DNA (eDNA) from both triatomine bugs and *Trypanosoma cruzi* for use in household surveillance in eastern Colombia, an endemic region for Chagas disease. Study findings demonstrated that FTA cards are a sensitive tool for detection of *Rhodnius prolixus* eDNA at temperatures between 21-32°C, deposited by individual, recently blood-fed nymphs. Additionally, cotton-tipped swabs are a relatively sensitive tool for field sampling of both *T. cruzi* and *R. prolixus* eDNA in infested households and are arguably more feasible due to their lower cost. eDNA detection should not yet replace current surveillance tools, but instead be evaluated in parallel as a more sensitive, higher-throughput, lower cost alternative. eDNA collection requires virtually no skills or resources *in situ* and therefore has the potential to be implemented in local endemic communities as part of citizen science initiatives to control Chagas disease transmission.

## Background

Chagas disease remains the most important parasitic infection in Latin America, responsible for the loss of 275,000 disability-adjusted life years (DALYs) in 2019 [1]. The geographical range of the aetiological agent, *Trypanosoma cruzi*, extends from the southern USA to Argentinean Patagonia, where it is transmitted by more than 100 species of triatomine bugs (Hemiptera: Reduviidae: Triatominae) to at least eight orders of domestic, synanthropic and sylvatic mammalian hosts [2, 3]. Human disease occurs when infected triatomine faeces enter through intact mucosa or abraded skin, causing an initial asymptomatic or non-specific self-limiting febrile illness, followed by life-long infection and potentially fatal cardiomyopathy (30-40% of infected individuals) [4]. Without a vaccine or highly efficacious treatment options for adults [5, 6], accurate detection of triatomine-infested houses and residual insecticide spraying of domestic and peri-domestic structures are crucial to prevent new cases [7-9]. Domiciliary populations of triatomine bugs have been successfully controlled by spraying of pyrethroid insecticides across large parts of Latin America, contributing to reductions in their distribution from an estimated 6.28 million km^2^ in the 1960s to less than 1 million km^2^ today [10]. Despite these achievements, the success of contemporary Chagas disease vector control programmes is threatened by persistent peri-domestic foci [11, 12], the emergence of insecticide resistance [13-15] and household invasion from sylvatic triatomine bug populations [16-19].

Chagas disease vector surveillance typically relies on identification of infested houses using timed-manual collections (TMCs), conducted by skilled personnel, with or without a dislodging spray [20, 21]. However, this methodology is laborious and suffers from several other drawbacks, including lack of standardization, substantial operational costs and limited sensitivity, especially when infestation is highly focal and/or triatomine bug densities are very low after a recent insecticide spraying campaign [22-24]. Community-based bug collections or bug notifications (with or without proof of collection) performed by householders have reported similar or sometimes superior levels of sensitivity to TMCs [25-27], but are prone to variability due to changes in motivation and skills of local voluntary residents [22, 25]. Additional passive devices, including double-sided sticky traps and artificial shelter units, often supplemented with semiochemical attractants, have shown more promising results in some endemic communities [28-30]. Following triatomine bug collection, further incrimination of risk of *T. cruzi* infection requires triatomine hindgut dissection and microscopic parasite visualization, parasite isolation by haemoculturing, or direct DNA extraction from faeces and PCR-based detection methods [31]. In some remote endemic areas, these procedures are not feasible, resulting in an inadequate understanding of triatomine bug infection rates.

Environmental DNA (eDNA) refers to genetic material sampled from the environment rather than the organism itself [32]. This technique can offer a non-invasive, highly sensitive alternative for surveillance, particularly of low levels of target organisms or new invasive species found in complex environments (e.g. water, soil or air) [33-35]. However, the sensitivity of eDNA detection is influenced by changes in temperature, rate of DNA degradation, target species density and ultraviolet light exposure [36-38]. Flinders Technology Associates (FTA®) cards consist of filter paper impregnated with a proprietary chemical mixture that lyses cells, inhibits overgrowth of bacteria and other microorganisms, denatures proteins and immobilises nucleic acids in a matrix, designed for long-term storage at room temperature [39]. In the context of vector-borne diseases, FTA cards have been used for successful preservation of different pathogens, including arboviruses [40-43], *Plasmodium falciparum, P. vivax* and *P. berghei* [40, 44, 45], *Mansonella ozzardi* [46] *Theileria pava* [47] and *Trypanosoma brucei* s.l. [48], as well as infected vector species, such as *Aedes* (*Ae*.) *aegypti* [49], *Ae. albopictus* [50], *Culex* species [51] and *Forcipomyia* (*Lasiohelea*) [52]. To date, the feasibility of using FTA cards for detection of triatomine bugs or *T. cruzi* has not been assessed. Therefore, this study evaluated the use of FTA cards and cotton-tipped swabs as low-technology, cost-effective tools for simultaneous surveillance of triatomine bug and *T. cruzi* eDNA in the laboratory and field.

## Methods

### Study design

This study had two components: (1) laboratory evaluation and optimisation of FTA cards to detect triatomine eDNA; (2) detection of triatomine and *T. cruzi* eDNA in household samples from an endemic region in eastern Colombia.

### Triatomine colony maintenance

The insectary at the London School of Hygiene and Tropical Medicine (LSHTM) provided the *R. prolixus* for this study. They were maintained at 25°C ± 60-80% relative humidity with 12 hours:12 hours light:dark cycles. This colony was derived from material sent to LSHTM from Venezuela in 1927. All *R. prolixus* individuals used in this study were 3^rd^ or 4^th^ instar nymphs. During the study period, triatomines were blood-fed with equine blood warmed through a Hemotek® feeder, used for experimentation, returned to the colony, and then re-used for experimentation once every 6 weeks.

### Optimising triatomine eDNA sampling

Before altering any environmental variables, which might have affected triatomine eDNA detection, an initial experiment was conducted to determine the optimal time for eDNA sampling. One hundred and five 3^rd^/4^th^ instar *R. prolixus* were blood-fed then immediately placed in different glass jars (dimensions: opening diameter 6.5cm; base diameter 9.5cm; height 14cm) in groups of 5 under standard insectary conditions (25.1-26.2°C, 68-74% relative humidity). For eDNA sampling, QIAcard® FTA® classic cards (Qiagen, UK) were used. These are cards treated with FTA®, a chemical which causes cell lysis and DNA immobilisation to isolate pure DNA. These were stapled to plain A4 paper and secured over the jar openings with an elastic band (Figure 1A). The jar was then inverted so that the *R. prolixus* were walking, urinating, and defecating on the card. Triatomines in the 21 jars were left on the FTA cards for 7 time points: 15 minutes, 30 minutes, 1 hour, 2 hours, 5 hours, 12 hours and 24 hours (Figure 1E). Each time point was trialled in biological triplicate, i.e. three independent jars were used per time point. After each time point, the glass jars were reverted (Figure 1C), *R. prolixus* returned to the colony, and the FTA cards removed and stored at - 20°C. eDNA extraction, amplification, and detection (described below) indicated that 24 hours was the optimal time point to leave the triatomines on the FTA cards to detect their eDNA. This time point was used in subsequent experiments.

**Figure 1.**
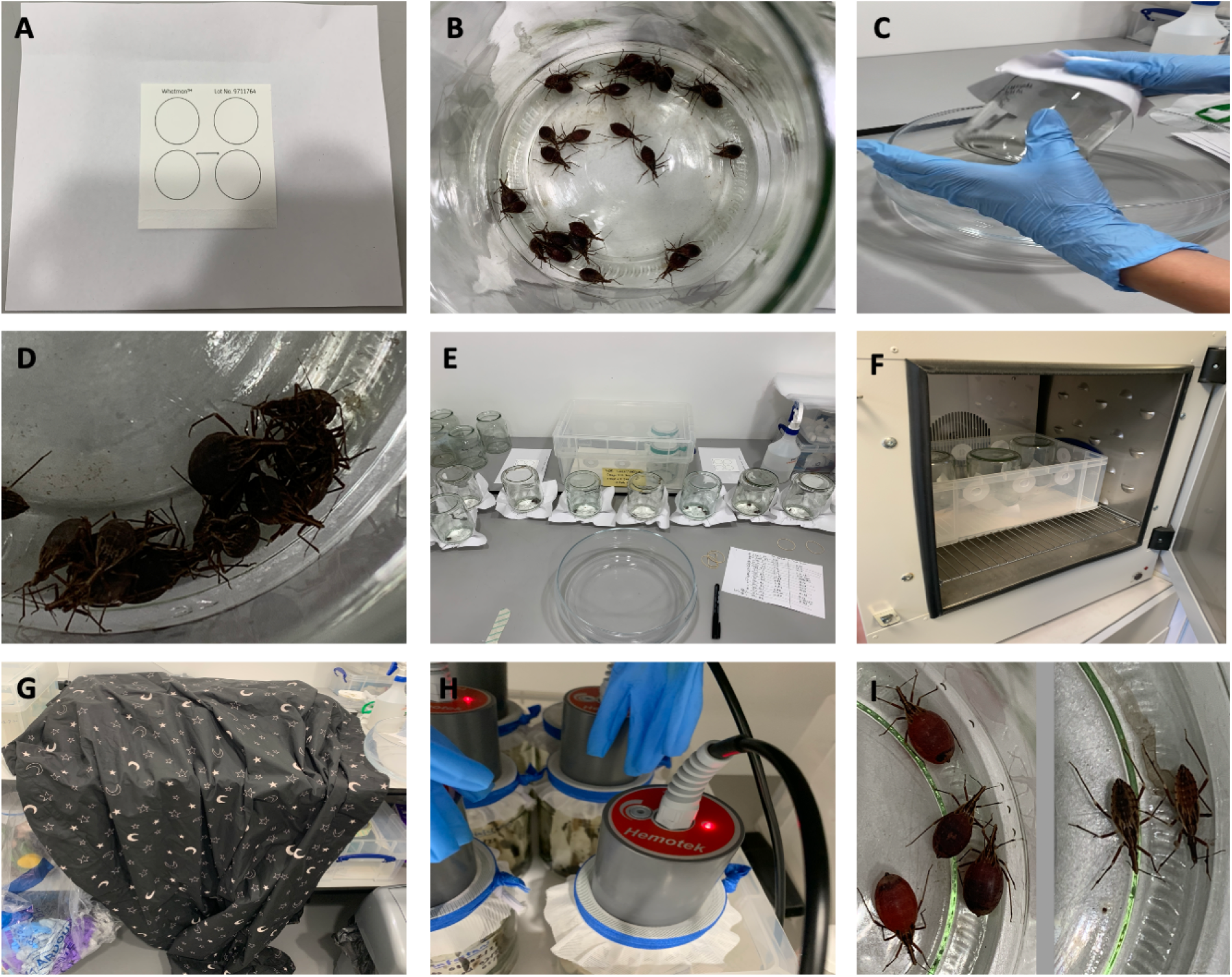
Photos of experimental set-ups. A: QIAcard® FTA® classic card, stapled to plain A4 paper, to be secured over a glass jar opening with an elastic band; B: bug number experiment with 25 blood-fed 3^rd^/4^th^ instar *R. prolixus*; C: process to invert glass jars; D: aggregating behaviour of 3^rd^/4^th^ instar *R. prolixus*; E: time point assay experiment; F: temperature experiment with incubator set to 32°C for 24 hours; G: darkness experiment with blackout blanket over inverted jars; H: blood-feeding the triatomines with Hemotek® feeder using equine blood; I: appearance of fed (left) *vs*. unfed bugs (right) in feeding status experiment.

### Altering triatomine eDNA environmental variables

Next a series of experiments investigating the impact of five different environmental variables (darkness, triatomine number, temperature, feeding status and degradation at ambient temperature) on triatomine eDNA detection were performed (Figure 2). All experiments were performed in biological triplicate. Negative controls were run concurrently in all experiments by placing FTA cards over empty glass jars. Temperature and humidity were consistently controlled in all experiments, except when temperature was the independent variable under evaluation. All experiments were performed consistently in 24 hours of artificial light, except when light was the independent variable under evaluation.

**Figure 2.**
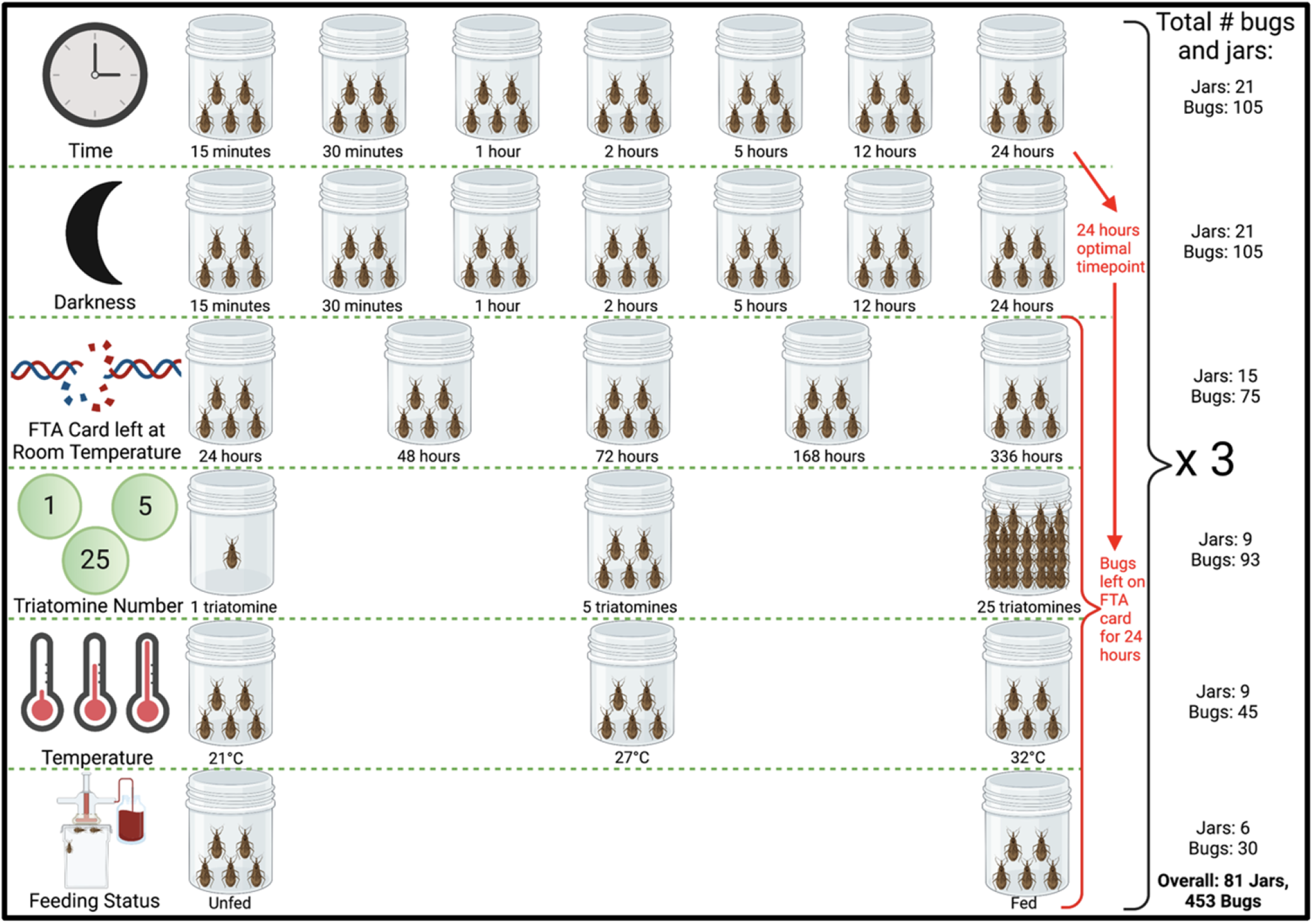
Design of experiments to investigate the impact of different environmental conditions on *R. prolixus* eDNA detection, including variables altered, number of jars and number of triatomines used.

### 1. Darkness

After blood-feeding 105 3^rd^/4^th^ instar *R. prolixus* and placing 5 in each jar, the jars were inverted onto FTA cards for one time point per group: 15 minutes, 30 minutes, 1 hour, 2 hours, 5 hours, 12 hours, and 24 hours, and covered by a blackout blanket to measure the effect of darkness on the quantity of eDNA sampled (Figure 1G and 2). Using all time points allowed us to compare whether the triatomines, which are nocturnal, would be more active in the dark than in artificial light. At the end of each time limit, bugs were returned to the colony and the FTA cards stored at -20°C.

### 2. Triatomine number

After blood-feeding 93 3^rd^/4^th^ instar *R. prolixus* and placing the following numbers in each glass jar: 1, 5 and 25 (Figure 1B, 1D, 1H and 2), the jars were inverted onto FTA card for 24 hours. After this time, bugs were returned to the colony and FTA cards stored at -20°C.

### 3. Temperature

After blood-feeding 45 3^rd^/4^th^ instar *R. prolixus* and placing 5 in each glass jar, the jars were inverted onto FTA cards for 24 hours. Each of the jars were placed in different temperatures in incubators for this duration: 21°C, 27°C, and 32°C (Figure 1F and 2). After 24 hours, bugs were returned to the colony and the FTA cards stored at -20°C.

### 4. Feeding status

Fifteen 3^rd^/4^th^ instar *R. prolixus* that had not blood-fed for 19 weeks were removed from the colony and 5 placed into each jar. An FTA card was secured to the opening and the jar inverted, and the bugs left for 24 hours. The same was done with 15 3^rd^/4^th^ instar *R. prolixus* which had been fed just before the FTA card was secured to the jar (Figure 1H, 1I and 2). After 24 hours, bugs were returned to the colony and the FTA cards were stored at -20°C.

### 5. Degradation at ambient temperature

After blood-feeding 105 3^rd^/4^th^ instar *R. prolixus* and placing 5 in each jar, the jars were inverted onto FTA card for 24 hours. After this point, bugs were returned to the colony and FTA cards detached from jars and left at room temperature for the following time points: 24 hours, 48 hours, 72 hours, 1 week, 2 weeks and 8 weeks to measure the effect of time at ambient conditions on eDNA degradation (Figure 2). Ambient temperature and humidity were recorded hourly using an EL-USB-2 RH/temperature data logger (Lascar Electronics, UK). The FTA cards were then stored at -20°C until eDNA extraction.

### Triatomine eDNA extraction from laboratory specimens

FTA cards from each different condition were cut in half and then into 1 × 2 cm strips using scissors sterilised between samples with 10% (v/v) bleach and 70% (v/v) ethanol. Two independent eDNA extractions were performed per FTA card. Strips from each half FTA card were placed into individual sterile 50 ml Falcon tubes (Fisher Scientific, UK), immersed in 5400μl ATL buffer and 600μl proteinase K (Qiagen, UK) and incubated overnight at 56°C. eDNA was extracted from 6 ml of sample using Qiagen DNeasy 96 Blood and Tissue kits (Qiagen, UK), according to the manufacturer’s protocol.

### Triatomine eDNA detection from laboratory specimens

Triatomine eDNA was detected using qPCR to amplify a fragment of the *R. prolixus 12S rRNA* gene. A standard curve of Ct values for this assay was generated using a 10-fold serial dilution of control *R. prolixus* gDNA (extracted from individual colony adults), to assess PCR efficiency. Genomic DNA concentration was determined using the Qubit 4 fluorometer 1X dsDNA HS assay (Invitrogen, UK).

Standard curve reactions were performed in a final volume of 10μl containing 2X PrimeTime® Gene Expression Master Mix (IDT, USA), 250nM of forward (P2B 5’-AAAGAATTTCCGGGTAATTTAGTCT-3’) and reverse (P6R 5’-GCTGCACCTTGACCTGACATT-3’) primers, 150nM of Triat probe (5’-/56FAM/TCAGAGGAA/ZEN/CCTGCCCTGTA/3IABkFQ/-3’) and 2μl genomic DNA (adapted from [53]). Reactions were run on a Stratagene Mx3005P Real-Time PCR system (Agilent Technologies, UK) at 95°C for 3 minutes, followed by 40 cycles of 95°C for 15 seconds and 60°C for 1 minute. All assays were run in technical triplicate alongside PCR no-template controls (NTCs).

To confirm primer and probe specificity to *R. prolixus*, the TaqMan assay was used to assess amplification of stock gDNA from *Triatoma (T*.*) infestans, T. dimidiata, Ae. aegypti, Ae. albopictus, Cx. quinquefasciatus, Anopheles (An*.*) arabiensis, An. gambiae* s.s., *An. coluzzii, An. stephensi, An. funestus* s.s., *Lutzomyia (L*.*) longipalpis* and *Phlebotomus (P*.*) papatasi*, using the same reaction conditions as the standard curve experiment.

eDNA detection in laboratory samples was performed in a final volume of 10μl containing 2X PrimeTime® Gene Expression Master Mix (IDT, USA), 250nM of forward and reverse primers, 150nM of probe and 4.35μl eDNA. Reactions were run on a Stratagene Mx3005P Real-Time PCR system (Agilent Technologies, UK) at 95°C for 3 minutes, followed by 40 cycles of 95°C for 15 seconds and 60°C for 1 minute. All assays were run in technical triplicate alongside PCR NTCs and *R. prolixus* positive controls.

### Household eDNA sampling and extraction

Due to the SARS-CoV-2 pandemic, we were unable to ship FTA cards to field collaborators in Colombia. Instead, between June-September 2021, cotton-tipped swabs (Guangzhou Improve Medical Instruments Co., Ltd, China) were used to sample triatomine and *T. cruzi* eDNA from different surfaces in houses and peri-domestic structures in Agualinda and San Isidro, municipality of Pore, Department of Casanare, Colombia (endemic for Chagas disease) and Prado Veraniego, municipality of Bogotá (non-endemic for Chagas disease) (Figure 3; Supplementary file S1). Householders completed a short questionnaire to confirm triatomine infestation and *T. cruzi* infection. Individual cotton swabs were packaged in small plastic bags filled with silica gel and shipped to the LSHTM at room temperature. In addition, a series of negative control cotton-tipped swabs, collected by sampling house walls in the United Kingdom, were prepared and processed in parallel to field specimens.

**Figure 3.**
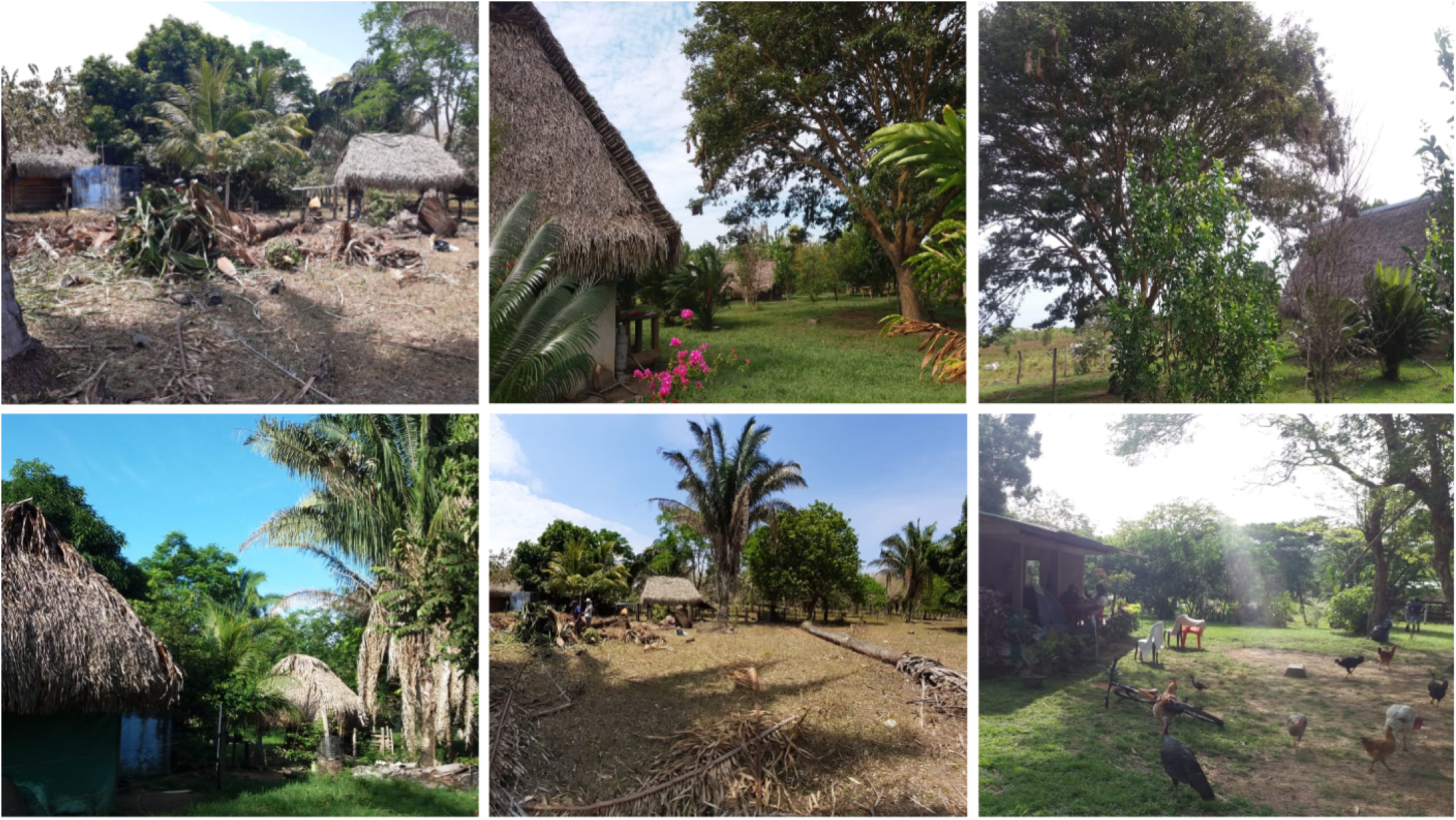
Houses and peri-domestic sites where eDNA was collected in Casanare Department, Colombia.

To extract eDNA, swabs were placed into individual sterile 50 ml Falcon tubes (Fisher Scientific, UK), immersed in 900μl ATL buffer and 100μl proteinase K (Qiagen, UK) and incubated overnight at 56°C. eDNA was extracted from the entire sample using Qiagen DNeasy 96 Blood and Tissue kits (Qiagen, UK), according to the manufacturer’s protocol.

### Triatomine and T. cruzi eDNA detection from field specimens

Parasite eDNA was detected using qPCR to amplify a fragment of the *T. cruzi* satellite DNA [54]. A standard curve of Ct values for this assay was generated using a 10-fold serial dilution of control *T. cruzi* gDNA (strain SMA6: TcI [55]), to assess PCR efficiency. Genomic DNA concentration was determined using the Qubit 4 fluorometer 1X dsDNA HS assay (Invitrogen, UK).

Standard curve reactions for *T. cruzi* qPCR were performed in a final volume of 10μl containing 2X PrimeTime® Gene Expression Master Mix (IDT, USA), 250nM of forward (Cruzi 1 5’-ASTCGGCTGATCGTTTTCGA-3’) and reverse (Cruzi 2 5’-AATTCCTCCAAGCAGCGGATA-3’) primers, 150nM of Cruzi 3 probe (5’-/5HEX/TTGGTGTCC/ZEN/AGTGTGTG/3IABkFQ-3’) and 2μl genomic DNA. Reactions were run on a Stratagene Mx3005P Real-Time PCR system (Agilent Technologies, UK) at 95°C for 3 minutes, followed by 40 cycles of 95°C for 15 seconds and 60°C for 1 minute. All assays were run in technical triplicate alongside PCR NTCs.

Simultaneous detection of *R. prolixus* and *T. cruzi* eDNA in field samples was performed in a final volume of 10μl containing 2X PrimeTime® Gene Expression Master Mix (IDT, USA), 250nM of P2B, 250nM of P6R, 150nM of Triat probe, 250nM of Cruzi 1, 250nM of Cruzi 2, 150nM of Cruzi 3 probe and 3.65μl eDNA. Reactions were run on a Stratagene Mx3005P Real-Time PCR system (Agilent Technologies, UK) at 95°C for 3 minutes, followed by 40 cycles of 95°C for 15 seconds and 60°C for 1 minute. All assays were run in technical triplicate alongside PCR NTCs and *R. prolixus* and *T. cruzi* positive controls.

### Data analysis

Stratagene MxPro qPCR software (Agilent Technologies, UK) was used to produce qPCR standard curves. qPCR assay limits of detection (LoD) and limits of quantification (LoQ) were determined using the “Generic qPCR LoD / LoQ calculator” [33], implemented in R version 4.0.2 [56]. All other statistical analyses were conducted in GraphPad Prism v9.2.0.

## Results

### R. prolixus and T. cruzi gDNA LoD / LoQ

To estimate the analytical sensitivity, linearity and dynamic range of the two qPCR assays for eDNA detection, serial dilutions of *R. prolixus* and *T. cruzi* gDNA were tested in three independent assays, with technical triplicates per assay. Both qPCR assays produced good linearity (*R*^2^=0.98 and 0.99 for *R. prolixus* and *T. cruzi* assays, respectively) and efficiencies of 90.94% and 93.07%, respectively (Figure 4). The LOD/LOQs were determined to be 0.015 and 0.0071 copies/reaction for *R. prolixus* and *T. cruzi* assays, respectively (Figure 4). The *R. prolixus* probe was highly specific to triatomine DNA; no amplification was detected with gDNA from other vector species: *Ae. aegypti, Ae. albopictus, Cx. quinquefasciatus, An. arabiensis, An. gambiae* s.s., *An. coluzzii, An. stephensi, An. funestus* s.s., *L. longipalpis* and *P. papatasi*. Low levels of cross-reactivity (i.e. high Ct values) were observed for *T. infestans* and *T. dimidiate* gDNA.

**Figure 4.**
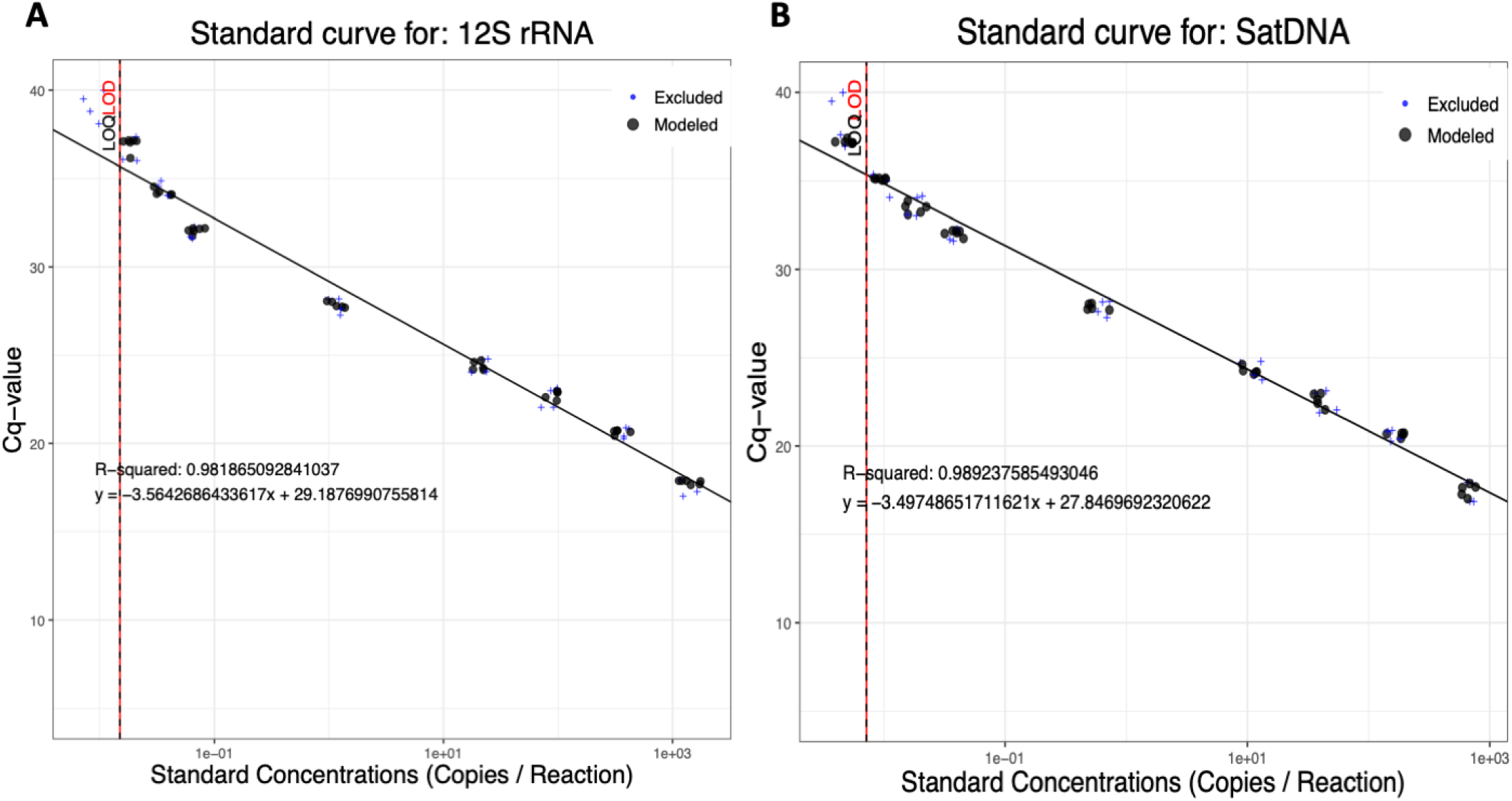
Standard curves for *R. prolixus* 12S rRNA (A) and *T. cruzi* satellite DNA (B) across serial gDNA dilution series.

### Triatomine eDNA environmental variables

The effects of altering five different environmental variables (darkness, triatomine number, temperature, feeding status and degradation at ambient temperature) on detection of *R. prolixus* eDNA from FTA cards were investigated (Figure 5). All qPCR sample data are reported in Supplementary file S2.

**Figure 5.**
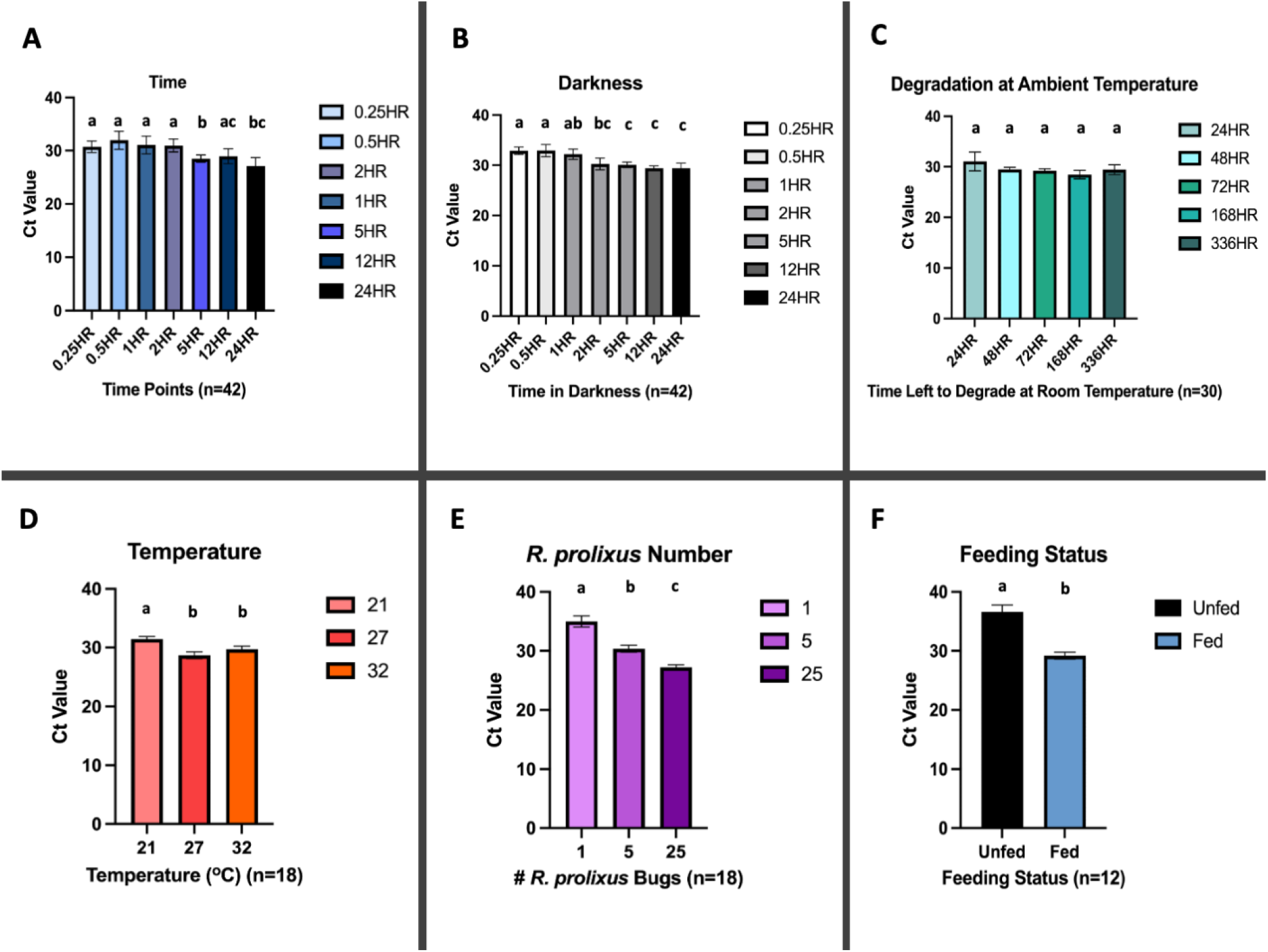
Detection of *R. prolixus* eDNA from FTA cards under different environmental conditions. A: time; B: darkness; C: degradation at ambient temperature; D: temperature; E: *R. prolixus* number; F: feeding status. N=number of separate DNA extractions performed. qPCR detection for all extractions were run in technical triplicate. Conditions sharing a superscript do not differ significantly (*p*>0.05). Error bars indicate 95% confidence intervals (CIs).

In artificial light, varying the amount of time *R. prolixus* 3^rd^/4^th^ instars had to walk/urinate/defecate on FTA cards from 15 minutes to 24 hours, did not substantially affect eDNA detection levels, with qPCR amplification observed at all timepoints (Figure 5A). In general, with increasing time, Ct values decreased significantly (Figure 5A). The average Ct values for each time point were 15 minutes: 30.72 [95% CI: 29.66-31.78]; 30 minutes: 31.96 [95% CI: 30.25-33.67]; 1 hour: 31.05 [95% CI: 29.40-32.70]; 2 hours: 30.97 [95% CI: 29.74-32.19]; 5 hours: 28.49 [95% CI: 27.80-29.17]; 12 hours: 28.94 [95% CI: 27.54-30.35]; and 24 hours: 27.10 [95% CI: 25.52-28.68]. Results from a parallel experiment, conducted in darkness, demonstrated comparable Ct values (Figure 5B), indicating that light/darkness did not impact *R. prolixus* eDNA detection. The average Ct values for the darkness experiment were 15 minutes: 32.89 [95% CI: 32.16-33.61]; 30 minutes: 32.92 [95% CI: 31.72-34.12]; 1 hour: 32.20 [95% CI: 31.17-33.23]; 2 hours: 30.26 [95% CI: 29.09-31.42]; 5 hours: 30.08 [95% CI: 29.52-30.64]; 12 hours: 29.39 [95% CI: 28.94-29.85]; and 24 hours: 29.37 [95% CI: 28.34-30.41]. With the lowest Ct values, 24 hours was chosen for subsequent experiments in which other environmental variables were altered.

A series of FTA cards were held at ambient temperature (mean temperature of 19.6°C [95% CI: 19.52-19.71°C] and mean relative humidity of 62.3% [95% CI: 62.08-62.48%]), once *R. prolixus* 3^rd^/4^th^ instars had walked/urinated/defecated on them for 24 hours, to determine how long triatomine eDNA takes to potentially degrade. *R. prolixus* eDNA was still detectable after 2 weeks, with no significant decline in detection level (Figure 5C). The average Ct values were 24 hours: 31.05 [95% CI: 29.18-32.92]; 48 hours: 29.44 [95% CI: 29.00-29.87]; 72 hours: 29.21 [95% CI: 28.82-29.59]; 1 week (168 hours): 28.44 [95% CI: 27.60-29.28]; and 2 weeks (336 hours): 29.40 [95% CI: 28.44-30.37]. However, by 8 weeks *R. prolixus* eDNA was no longer detectable (all six DNA extractions failed to amplify during all three technical qPCR replicates).

To simulate a range of household temperatures in Chagas endemic regions, *R. prolixus* 3^rd^/4^th^ instars were allowed to walk/urinate/defecate on FTA cards for 24 hours at 21°C, 27°C and 32°C (Figure 5D), with significantly lower Ct values detected at 27°C (average Ct value: 28.69 [95% CI: 28.11-29.27]) and 32°C (average Ct value: 29.72 [95% CI: 29.20-30.24]), compared to 21°C (average Ct value: 31.45 [95% CI: 31.03-31.88]) (one-way ANOVA, F(2, 45) = 29.89, *p*<0.0001).

A strong inverse relationship between number of *R. prolixus* 3^rd^/4^th^ instars allowed to walk/urinate/defecate on FTA cards for 24 hours was also apparent (Figure 5E). The average Ct values for this experiment were 1 bug: 34.99 [95% CI: 34.06-35.92]; 5 bugs: 30.36 [95% CI: 29.77-30.95]; and 25 bugs: 27.22 [95% CI: 26.77-27.66], with significantly greater levels of detection with increasing triatomine density (one-way ANOVA, F(2, 50) = 148.2, *p*<0.0001).

Finally, *R. prolixus* physiological status also had a significant impact on eDNA detection, with significantly higher levels of sensitivity observed for recently blood-fed bugs (Figure 5F). The average Ct values were unfed: 36.59 [95% CI: 35.39-37.80] and fed: 29.20 [95% CI: 28.60-29.79] (t-test, *p*<0.0001). Furthermore, the rate of qPCR non-amplification was higher among the unfed bug group, with three out of six DNA extractions failing to amplify during all three technical replicates; one extraction amplified once, and two extractions amplified twice.

### Triatomine and T. cruzi eDNA detection from field specimens

In Colombia, sixty-three houses were sampled in Prado Veraniego (n=30), Bogotá (non-endemic for Chagas disease), and Agualinda (n=14) and San Isidro (n=19), Casanare (endemic for Chagas disease), for *R. prolixus/T. cruzi* eDNA using cotton-tipped swabs. Samples were collected from a range of domestic and peridomestic locations, including from living room walls, living room ceilings, TV room walls, kitchen walls, dining room walls, stables and hen nests (Supplementary file S1). All houses in Agualinda and San Isidro were confirmed as infested with triatomine bugs by self-reporting householders, while all houses in Prado Veraniego were reported as uninfested. Only two households in San Isidro reported human *T. cruzi* infection.

Among 33 households in Agualinda and San Isidro with confirmed triatomine infestations, *R. prolixus* eDNA was detected in 20, giving an estimated sensitivity of 60.6% (Figure 6); average Ct value was 29.56 [95% CI: 28.20-30.91]. In these same houses, *T. cruzi* eDNA was detected in 31, including the two houses with reported human infections (houses #26 and 28; Figure 6); average Ct value was 34.17 [95% CI: 33.56-34.78]. Among 30 households in Prado Veraniego, which were negative for triatomine infestation, no *R. prolixus* or *T. cruzi* eDNA was detected, giving an estimated specificity of 100%. Finally, no *R. prolixus* or *T. cruzi* eDNA was amplified from 15 cotton-tipped swabs sampled from walls in the UK.

**Figure 6.**
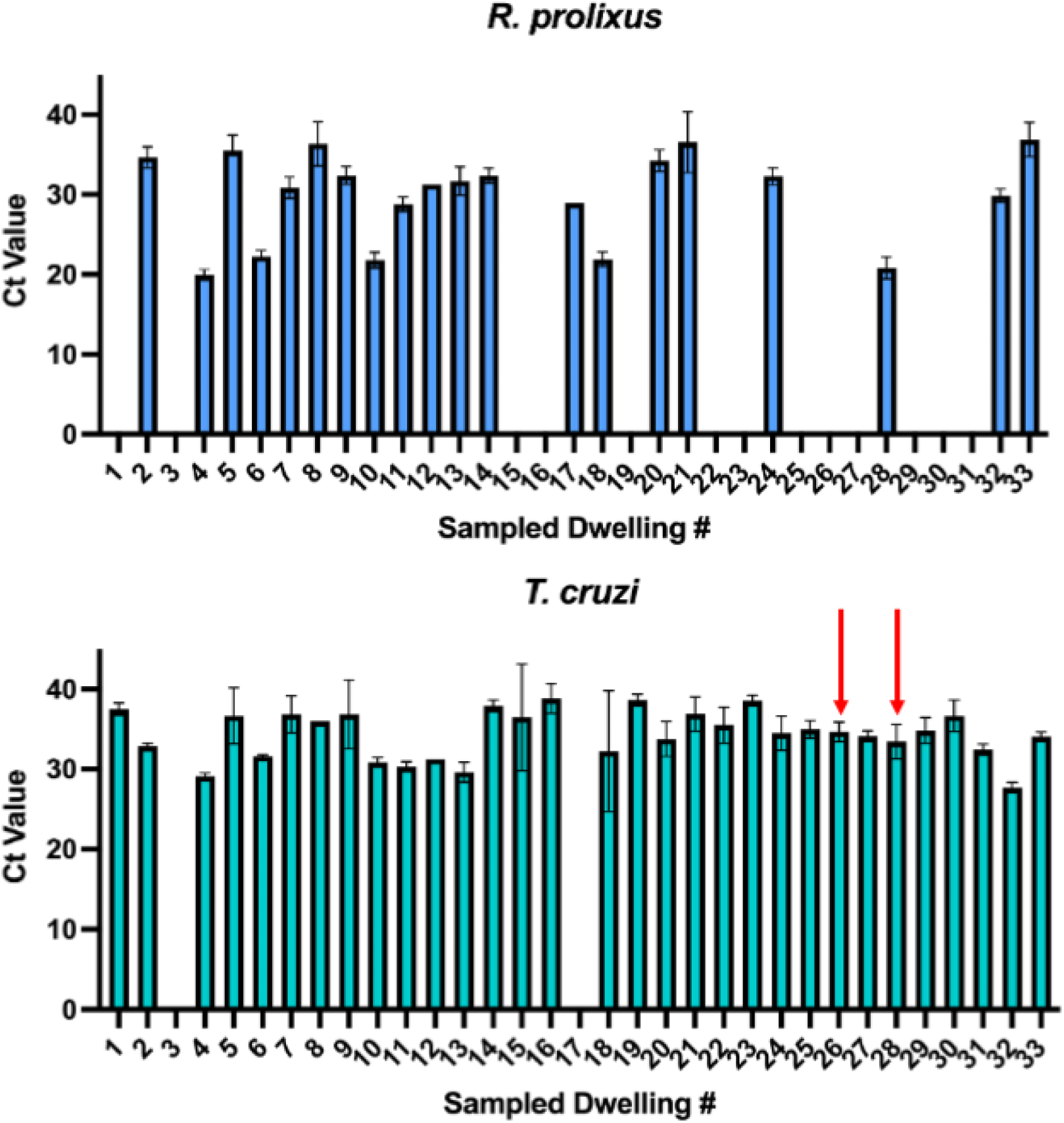
Detection of *R. prolixus* and *T. cruzi* eDNA from cotton swabs taken from 33 house with confirmed triatomine infestations in Casanare, Colombia. Red arrows indicate houses with reported human *T. cruzi* infection.

## Discussion

Accurate surveillance of triatomine household infestation is crucial for Chagas disease vector control. However, no gold standard vector detection method with high levels of sensitivity or specificity is currently available. Given there are several intrinsic features of triatomine bug behaviour and the lifecycle of *T. cruzi* which lead to deposition of pathogen and vector eDNA in infested houses, this study evaluated the use of FTA cards or cotton-tipped swabs for simultaneous, non-invasive parasitological and entomological surveillance in eastern Colombia, an endemic region for Chagas disease.

In the proof-of-concept laboratory experiments, *R. prolixus* eDNA from five 3^rd^/4^th^ instar nymphs was successfully isolated and amplified from FTA cards after as little as 15 minutes of contact time under standard insectary conditions. This indicates that FTA cards may be able to detect transient triatomine movements and lighter triatomine infestations comprising of immature vector stages; these are features of new infestations and/or vector population recovery post insecticide spraying, and notoriously harder to measure with conventional TMCs [25]. While altering contact time (up to 24 hours), and light/dark cycles, did not significantly improve *R. prolixus* eDNA detection, increasing temperature from 21°C to 27-32°C and triatomine bug density from 1-25 bugs, were both inversely correlated with higher levels of qPCR amplification. Due to the laboratory conditions being highly artificial, the sensitivity of the FTA cards may have been overestimated by placing triatomines in smaller confined spaces than their colony jars and altering their light:dark cycles, causing them to move or defecate more due to stress. *R. prolixus* physiological status also impacted eDNA detection, with significantly higher levels of sensitivity observed for recently blood-fed bugs, compared to those starved for 19 weeks, suggesting that the majority of detectable eDNA is likely derived from triatomine faeces/urine compared to tarsal deposition upon contact. These findings are consistent with FTA card evaluations for other vectors and pathogens of public health importance, demonstrating direct dose responses between detection levels and target organism density [40, 42]. Under simulated conditions of degradation, *R. prolixus* eDNA was shown to be stable on FTA cards for at least two weeks at room temperature, which aligns with previous studies reporting successful pathogen isolation from cards stored for several weeks under similar conditions; published observations indicate longer-term stability can be achieved when FTA cards are stored at 4°C [39].

In the field specimens, *R. prolixus* eDNA was detected from infested houses with an estimated sensitivity of 60.6% which is comparable with reported detection levels for TMCs, householder collections and other trapping techniques [25, 28, 57]. Given that household triatomine presence was self-reported, one explanation for the number of false negative houses may be inaccuracies in resident reports, infestation with other local triatomine species (ten triatomine species, including *R. prolixus, Triatoma maculata* and *Panstrongylus geniculatus*, have been identified in Casanare [58]), relative sensitivity of using cotton-tipped swabs (these are not specifically optimised for DNA capture like FTA cards), or the LOD of our qPCR assay. More importantly, this technique was shown to be 100% specific in this particular field setting; false-positives in triatomine vector surveys arise due to taxonomic errors (i.e. mis-identifying non-triatomine reduviid nymphs or non-vector species), use of indirect proxies of vector infestation (e.g. triatomine bug faecal streaks which may be confused with those of other arthropods [59]) or when householders report vector presence without visual confirmation [22]. This observation highlights the potential for this technique to accurately identify foci of residual triatomine infestation, which are important sources of operational failure of current Chagas disease vector control programmes [60, 61].

Interestingly, *T. cruzi* eDNA was amplified from 93.9% of infested houses, when only 6.06% reported human infection. We excluded possible laboratory contamination as a confounder by processing known negative controls (cotton-tipped swabs wiped on house walls in the UK) in parallel with field specimens at every analytical step. Instead, this discordance may reflect differences in the gene copy number between the two targets used for qPCR detection; in our assay the LOD for the *T. cruzi* satellite DNA was an order of magnitude more sensitive compared to the *12S rRNA* in *R. prolixus*. Alternatively, these findings may be indicative of active infected vectors in these houses; parasite transmission is known to be highly precarious and inefficient, requiring an estimated 900-4000 infected contacts per case [62]. While human infection was only self-reported in two houses, *T. cruzi* transmission in this area is also under-diagnosed due to substantial heterogeneity in acute symptomology [58] and other significant barriers to adequate healthcare, including lack of diagnostics, infrastructure and financial investment, and limited physician awareness [63]. Regardless, the potential presence of residual parasite eDNA in these houses requires further investigation, including confirmation of infected vectors using a second entomological surveillance method and serodiagnosis of householders.

Findings from this study provide insights into the feasibility of using FTA cards or cotton-tipped swabs for community-level surveillance. Based on laboratory results, FTA cards could be taped to house walls for several weeks at temperatures between 21-32°C and potentially detect eDNA deposited by individual, recently blood-fed *R. prolixus* nymphs. Field specimens further confirmed our ability to amplify *R. prolixus/T. cruzi* eDNA deposited on house walls in Casanare using a cheaper, lower-technology tool. By comparison to TMCs and other trapping techniques, these methodologies require virtually no skills or training and do not involve residents actively exposing themselves to potentially infectious triatomines. This surveillance strategy could be integrated into newly developed citizen science initiatives for Chagas disease, which have used social media applications and behavioural design frameworks to improve community disease awareness and reporting of house infestation [64-66]. Additional longitudinal evaluations of this methodology are needed alongside parallel TMCs and community serosurveys, to quantitatively evaluate this technique, to establish true LOD of parasite/vector eDNA in the field using both FTA cards and cotton-tipped swabs, to determine how seasonal changes in triatomine population dynamics affect eDNA degradation and to optimise timing and logistics of eDNA wall sampling. While this study only investigated presence/absence of *T. cruzi* and *R. prolixus*, further research is warranted to assess the possibility of using recovered eDNA for community-wide blood-meal analysis, surveillance of molecular insecticide resistance and characterization of parasite and vector population genetic structures, including reinfestation dynamics after insecticide spraying campaigns [13, 17, 67]. These aspects are pivotal in the development of effective vector control programmes in Chagas disease endemic regions.

## Conclusions

This study validated the use of FTA cards and cotton-tipped swabs for simultaneous entomological and parasitological Chagas disease surveillance. Study findings demonstrated that FTA cards are a sensitive tool for detection of *R. prolixus* eDNA at temperatures between 21-32°C, deposited by individual, recently blood-fed 3^rd/^4^th^ instar nymphs. Additionally, cotton-tipped swabs are a relatively sensitive tool for field sampling of both *T. cruzi* and *R. prolixus* eDNA *in situ* and are arguably more feasible due to their lower cost. eDNA detection should not yet supplant current methods such as TMCs, but instead be evaluated alongside them as a more sensitive, higher-throughput, lower cost prospective alternative. eDNA collection can be implemented in local endemic communities as part of citizen science initiatives to monitor and control Chagas disease transmission. Further studies are needed to investigate the feasibility of using recovered eDNA for exploratory genetic analyses.

## Data Availability

All data produced in the present work are contained in the manuscript.

## List of abbreviations

Ct: cycle threshold
DALYs: disability-adjusted life years
eDNA: environmental deoxyribonucleic acid
FTA: Flinders Technology Associates
gDNA: genomic deoxyribonucleic acid
HS: high sensitivity
LoD: limits of detection
LoQ: limits of quantification
LSHTM: London School of Hygiene and Tropical Medicine
NTC: no-template controls
qPCR: quantitative polymerase chain reaction
rRNA: ribosomal ribonucleic acid
TMC: timed-manual collections

## Declarations

### Ethics approval and consent to participate

Ethical approval for the study was obtained from the London School of Hygiene and Tropical Medicine (LSHTM; ref#25638) and the Universidad del Rosario (“Genómica, evolución y biogeografía de especies del género *Rhodnius*: vectores de la enfermedad de Chagas*”* act number 007/2016) and all study procedures were performed in accordance with relevant guidelines and regulations.

### Consent for publication

Not applicable.

### Availability of data and materials

Not applicable.

### Competing interests

The authors declare that they have no competing interests.

### Funding

Study funding was provided by London School of Hygiene and Tropical Medicine MSc bench fees awarded to GG and a Wellcome Trust/Royal Society Sir Henry Dale Fellowship awarded to TW (101285/Z/13/Z): https://wellcome.org and https://royalsociety.org.

### Authors’ contributions

GG, PU, LBG, SB, JD and LAM designed the study. GG led the laboratory experiments and performed the molecular analysis, under the supervision of LAM. LBG and SB maintained the triatomine colonies and participated in data collection. PU and JD led the entomology field activities and participated in data collection. MK and TW provided laboratory resources and participated in data analysis and interpretation. LAM drafted the manuscript, which was revised by co-authors. All authors read and approved the final manuscript.

## Acknowledgements

The authors would like to thank Bethanie Pelloquin for assistance with DNA extraction.

## Notes

### Competing Interest Statement

The authors have declared no competing interest.

### Author Declarations

Ethical approval for the study was obtained from the London School of Hygiene and Tropical Medicine (LSHTM; ref#25638) and the Universidad del Rosario (Genomica, evolucion y biogeografia de especies del genero Rhodnius: vectores de la enfermedad de Chagas act number 007/2016) and all study procedures were performed in accordance with relevant guidelines and regulations.

